# On the track of the D839Y mutation in the SARS-CoV-2 Spike fusion peptide: emergence and geotemporal spread of a highly prevalent variant in Portugal

**DOI:** 10.1101/2020.08.10.20171884

**Authors:** Vítor Borges, Joana Isidro, Helena Cortes-Martins, Sílvia Duarte, Luís Vieira, Ricardo Leite, Isabel Gordo, Constantino P. Caetano, Baltazar Nunes, Regina Sá, Ana Oliveira, Raquel Guiomar, João Paulo Gomes, Portuguese network for SARS-CoV-2 genomics

## Abstract

Mutations in the Spike motif predicted to correspond to the fusion peptide are considered of interest as this domain is a potential target for anti-viral drug development that plays a pivotal role in inserting SARS-CoV-2 into human cell membranes. We tracked the temporal and geographical spread of a SARS-CoV-2 variant with the Spike D839Y mutation in the fusion peptide, which was detected early during the COVID-19 epidemic in Portugal. We show that this variant was most likely imported from Italy in mid-late February 2020, becoming prevalent in the Northern and Central regions of Portugal, where represented 22% and 59% of the sampled genomes, respectively, until the end of April 2020. Based on our high sequencing sampling during the early epidemics [15,5% (1275/8251) and 6,0% (1500/24987) of all confirmed cases until the end of March and April, respectively)], we estimate that, between March 14^th^ and April 9^th^ (covering the exponential phase of the epidemic), the relative frequency of Spike Y839 variant increased at a rate of 12.1% (6.1%-18.2%, CI 95%) at every three days, being potentially associated with one in each four (20.8-29.7%, CI 95%) COVID-19 cases in Portugal during the same period. This observation places the Spike Y839 variant in the origin of the largest SARS-CoV-2 transmission chain during the first month of the COVID-19 epidemic in Portugal. We hypothesize that population/epidemiological effects (founder effects) and enhanced selective advantage might have concomitantly contributed to the increasing frequency trajectory of the Spike Y839 variant. Screening of the D839Y mutation globally confirmed its detection in 12 additional countries, even though the huge differences in genome sampling between countries hampers any accurate estimate of D839Y global frequency. In summary, our data points out that SARS-CoV-2 Spike Y839 variants, namely the descendent variant of the globally spread G614 variant detected in Portugal, need continuous and close surveillance.

## Background

The causative agent of COVID-19, the novel coronavirus SARS-CoV-2 (Severe Acute Respiratory Syndrome Coronavirus 2), is a tremendous threat globally, already leading to more than 18 million confirmed cases and approximately 700 thousand deaths worldwide, as of August 6^th^, 2020 (https://www.who.int/emergencies/diseases/novel-coronavirus-2019/situation-reports/). SARS-CoV-2 is a single-stranded positive-sense RNA virus possessing a ~29900 bp genome (Wu et al, 2020; Zhou et al, 2020) that encodes four structural proteins: Spike (S), envelope (E), membrane (M) and nucleocapsid (N) (Tang et al, 2020). Spike protein governs the binding of SARS-CoV with its receptor angiotensin-converting enzyme 2 (ACE2) in human cells, the fusion between the viral and host cell membranes and, thus, the virus entry (Tang et al, 2020; Li W et al, 2020; Li L et al, 2020; Walls et al, 2020; Shang et al, 2020a; Shang et al, 2020b; Lan et al, 2020). Besides the need of an effective S receptor binding, infectivity of SARS-CoV-2 is also dependent on cleavage of the S protein by host proteases, activating it for fusion with the cell membrane (Tang et al, 2020; Shang et al, 2020a). The S protein, which decorates the virion surface, also induces neutralizing antibodies and it is therefore the key target for vaccine development (Tang et al, 2020; Walls et al, 2020; Du et al, 2020). In this context, it is of upmost importance to track the genetic diversity and evolution of circulating SARS-CoV-2 at regional and global levels, with special focus on detecting the emergence and monitoring the spread of S variants.

Although surveillance has been expectedly focused on genetic changes affecting the S receptor binding domain (RBD) within S1 subunit, as they may directly affect the binding to ACE2 and ultimately SARS-CoV-2 host range and infectivity/transmissibility (Tang et al, 2020; Wu et al, 2020; Zhou et al, 2020; Andersen et al, 2020), changes in other S protein domains should also be closely surveyed, particularly when they are linked to variant frequency increase at local, regional or global levels (Korber et al, 2020; Li et al, 2020). A variant carrying the Spike D614G mutation (affecting the S2 subunit) stands out as a remarkable example, as it became dominant worldwide during the first months of the pandemic (Nextstrain; GISAID; Korber et al, 2020) with recent studies suggesting that the G614 variant might be linked to an increased transmissibility but not pathogenicity (Korber et al, 2020). Spike amino acid 614 (within S2 subunit) is pocketed adjacent to the fusion peptide (FP), which is the functional fusogenic element of the S protein (Tang et al, 2020), and near the expected cleavage site, suggesting that G614 might have induced a conformational change in the Spike protein influencing the dynamics of the spatially proximal fusion peptide, thereby resulting in the altered infectivity (Korber et al, 2020). Hence, other mutations of interest outside the S protein RBD have been highlighted (Korber et al, 2020), particularly those falling within the Spike FP (predicted region 816-855) or fusion peptide proximal regions (Korber et al, 2020; Tang et al, 2020), due to the critical role of these motifs in inserting SARS-CoV-2 into the membrane of human cells (Tang et al, 2020; Cai et al, 2020). The fusion mechanism (governed by Spike S2 subunit) is also pointed out as an important target for the development of specific drugs against coronavirus, since it is expectedly less mutable than the surface-exposed and immunogenic S1 RBD, which, from an evolutionary point-of-view, is not an ideal target for broad-spectrum antiviral inhibitor development (Xia et al, 2019; Tang et al, 2020).

In the present study, we evaluated the temporal and geographical spread of a SARS-CoV-2 variant carrying the Spike protein amino acid change D839Y (within the fusion peptide) and show that this variant was introduced early in the epidemics in Portugal, becoming highly prevalent in the Northern and Central regions of Portugal (representing 22% and 59% of the sampled genomes until the end of April 2020, respectively). The emergence and high frequency of Spike Y839 variant (a descendent variant of the globally spread G614 variant) in SARS-CoV-2 circulating in Portugal, as well as the detection of the D839Y mutation in 12 additional countries (representing around 5% of sampled genomes in three countries), sustain that continuous molecular surveillance is pivotal to detect and monitor SARS-CoV-2 variants with biological/epidemiological relevance.

## Results

### Introduction and spread of the SARS-CoV-2 Spike Y839 variant in Portugal

Acting as the National Reference Laboratory for SARS-CoV-2, the Portuguese National Institute of Health (INSA) Doutor Ricardo Jorge rapidly established the genome-based molecular surveillance of SARS-CoV-2 in Portugal. A website (https://insaflu.insa.pt/covid19) was launched, providing updated data regarding the analysis of the SARS-CoV-2 genetic diversity and geotemporal spread. Also, "situation reports” with major highlights are sent periodically to participating laboratories, national and regional public health authorities and other stakeholders. As of July 23, 2020, INSA had analysed 1516 genome sequences (Table S1), enrolling 15.5% (1275/8251) and 6.0% (1500/24987) of all confirmed cases detected until the end of March and April, respectively. This very high sequencing sampling during the early epidemics allowed us to pursue one of the main objectives of the study, i.e., to identify the scenario of the introductions and early within-country dissemination of SARS-CoV-2 in Portugal (manuscript in preparation). The distribution of the 1516 SARS-CoV2 genomes from Portugal (https://microreact.org/project/nDGsJKFv7gQTj1q8CQwwKR/18a0a470) according to Nextstrain clade definition (https://clades.nextstrain.org/) follows, in general, the same trend observed at European level (https://nextstrain.org/ncov/europe). Most viruses (89.8%) integrate the phylogenetic branch enrolling clades 20A (40.8%), 20B (46.1%) and 20C (2.9%), carrying, among other genetic markers, the globally dispersed (Nextstrain; GISAID; Korber et al, 2020) nucleotide change A23403G, leading to the D614G amino acid replacement in the Spike protein. Clades 19A and 19B were found at the relative frequency of 7.4% and 2.8% in this dataset, respectively. Within clade 20A, a SARS-CoV-2 variant carrying the Spike amino acid change D839Y (due to a G24077T SNP) has been detected early (March 7^th^, 2020) during the COVID-19 epidemic in Portugal.

The Spike Y839 variant was most likely imported from Italy in mid-late February 2020, as all first detected genomes were collected from individuals from the Northern region of Portugal epidemiologically linked to infected individuals that returned from Milan in February 22^nd^-23^rd^. Concordantly, the first D839Y genome sequence reported worldwide was collected in Italy (Lombardy) on February 21^st^ (Italy/PV-5314-N/2020; GISAID accession number EPI_ISL_451307) (Table S2), being identical (i.e, Nextstrain clade 20A background plus the G24077T SNP) to the "founder” Spike Y839 variant genome sequences detected in Portugal (https://insaflu.insa.pt/covid19). Current data and ongoing epidemiological investigations do not support the existence of additional introductions of this variant. The Spike Y839 variant became particularly prevalent in Portugal, representing about 20% of all sampled genomes collected until the end of March (255/1275) or the end of April (287/1500) (Figure 1). Its circulation was particularly marked in Northern (epicenter of COVID-19 pandemic in Portugal) and Central regions of Portugal, representing 22% and 59% of the sampled genomes until the end of April 2020, respectively (Figure 2 and 3; https://microreact.org/proiect/nDGsJKFv7gQTi1q8CQwwKR/0489f840). In the same period, four districts (with at least 50 genomes sampled as of April 30^th^) revealed the highest Y839 relative frequency: Braga (16.3%), Porto (25.2%), Aveiro (68.1%) and Guarda (72.7%) (Figure S1). We estimate that the relative frequency of Y839 increased at a rate of 12.1% (6.1%-18.2%, CI 95%) every three days between March 14^th^ and April 9^th^, being potentially associated with 3793 (3177-4542, CI 95%) COVID-19 cases in Portugal during that period [representing 24.8% (20.8-29.7%, CI 95%) of the total confirmed cases reported in the same period) (Figure 4). Hence, our data supports that the Spike Y839 variant was circulating in Portugal since mid-late February (more than one week before the first COVID-19 confirmed case at March 2^nd^, 2020), being most likely responsible for the largest SARS-CoV-2 transmission chain occurred during the first 1-1.5 months of COVID-19 epidemic in Portugal. Noteworthy, the Spike Y839 variant is strongly linked to a large and worrying COVID-19 “local” outbreak occurring in a small municipality (Ovar) in the coastal side of the Central region of the country (District of Aveiro), with 80% of the genomes collected from this municipality carrying the Y839 variant. Ovar was the only municipality from Portugal mainland that was subjected to strict local quarantine and lockdown measures (from March 18^th^ to April 18^th^), presenting 692 confirmed cases (about 1250 cumulative cases per 100000 inhabitants) by the end of April. The Public Health Unit of Primary Care Cluster of Baixo Vouga (covering several municipalities, including Ovar), has been carrying out a deep investigation leading to the identification of clusters of potential epidemiologically linked confirmed cases (“epiclusters”) among 1556 monitored COVID-19 cases (by April 30^th^). Our SARS-Co V-2 genome collection (as of April 30^th^) includes representative samples of 41 of those potential epiclusters (covering a total of 420 confirmed cases), with 33 (323 confirmed cases) (77%) being exclusively associated with Spike Y839 variant (Figure S2). In a conservative manner, it is reasonable to extrapolate that Spike Y839 variant is potentially associated with about 1200 cases (77% of the total 1556 monitored cases) in the region covered by Primary Care Cluster of Baixo Vouga. Still, the Spike Y839 variant disseminated far beyond the coastal municipality of Ovar and neighborhood municipalities. In the inland region of the country, Y839 variant was for instance linked to a large cluster of infected individuals living/working in a nursing home in Vila Nova de Foz Côa municipality (District of Guarda) detected by the end of March, and more than 50% of Y839 genomes detected in April were collected in the District of Viseu (Figure S1; Figure S3; https://microreact.org/proiect/2kh3TRVYB9gWGRpNSJWDW5/b6c659e0), which borders the district of Aveiro. In total, the highly prevalent Spike Y839 variant was already detected in 44 municipalities across 11 out of the 18 Districts of Portugal mainland, consolidating that this descendent variant of the globally spread G614 variant had a remarkable weight in the early COVID-19 epidemic in Portugal.

**Figure 1.**
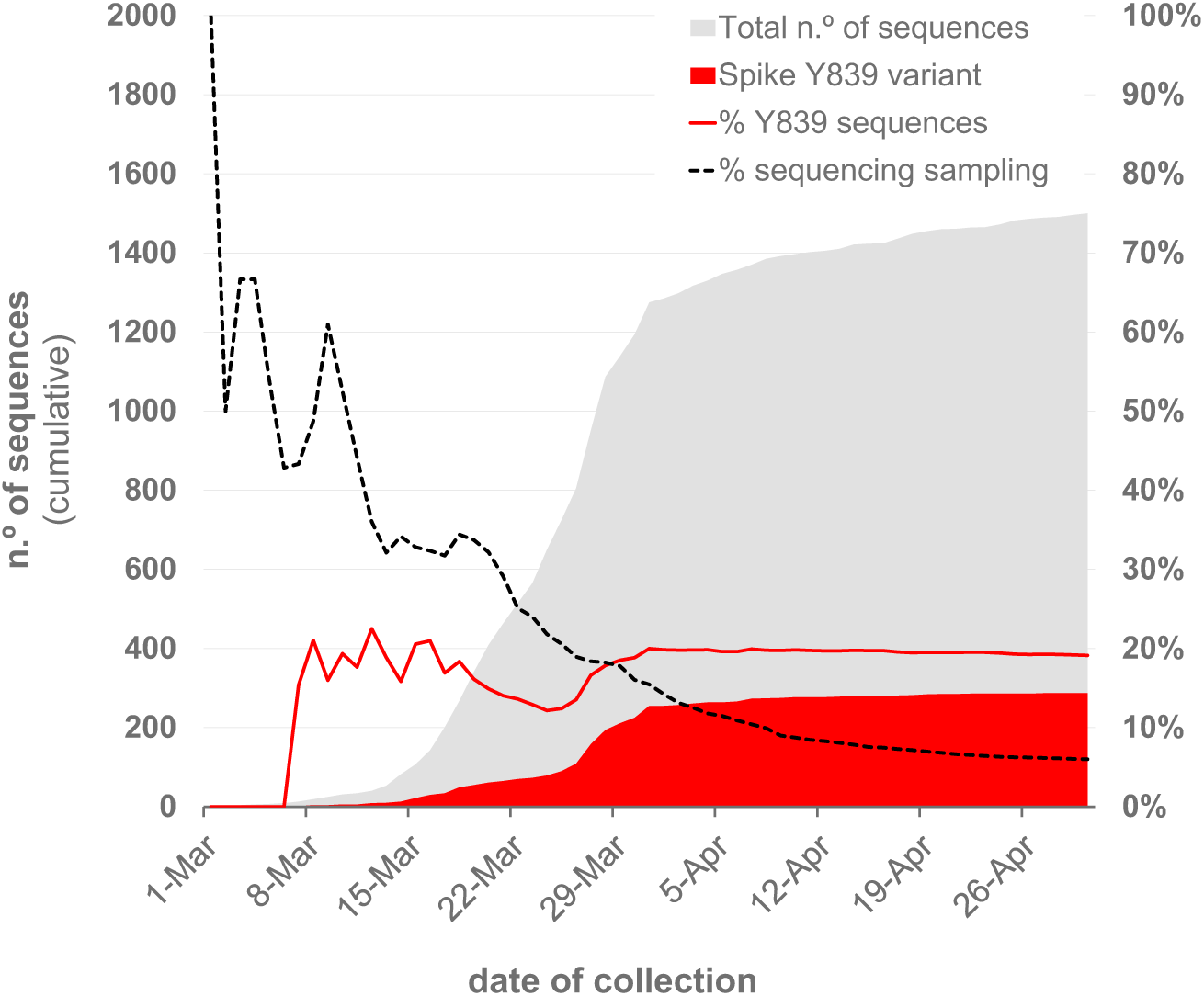
Overview of the SARS-CoV-2 genome sequencing sampling in Portugal and cumulative relative frequency of the circulating Spike Y839 variant, as of April 30^th^’ 2020 (n=1500) Area plots (left y-axis) reflect the cumulative total number of SARS-CoV-2 genome sequences (gray) and Spike Y839 variant sequences (red) obtained in Portugal during the first two months of the epidemic. Lines (right y-axis) display the cumulative percentage of COVID-19 confirmed cases for which SARS-CoV-2 genome data was generated (“sequencing sampling” - black dash line) and the cumulative proportion of the Spike Y839 variant sequences (red line) detected in Portugal during the same period.

**Figure 2.**
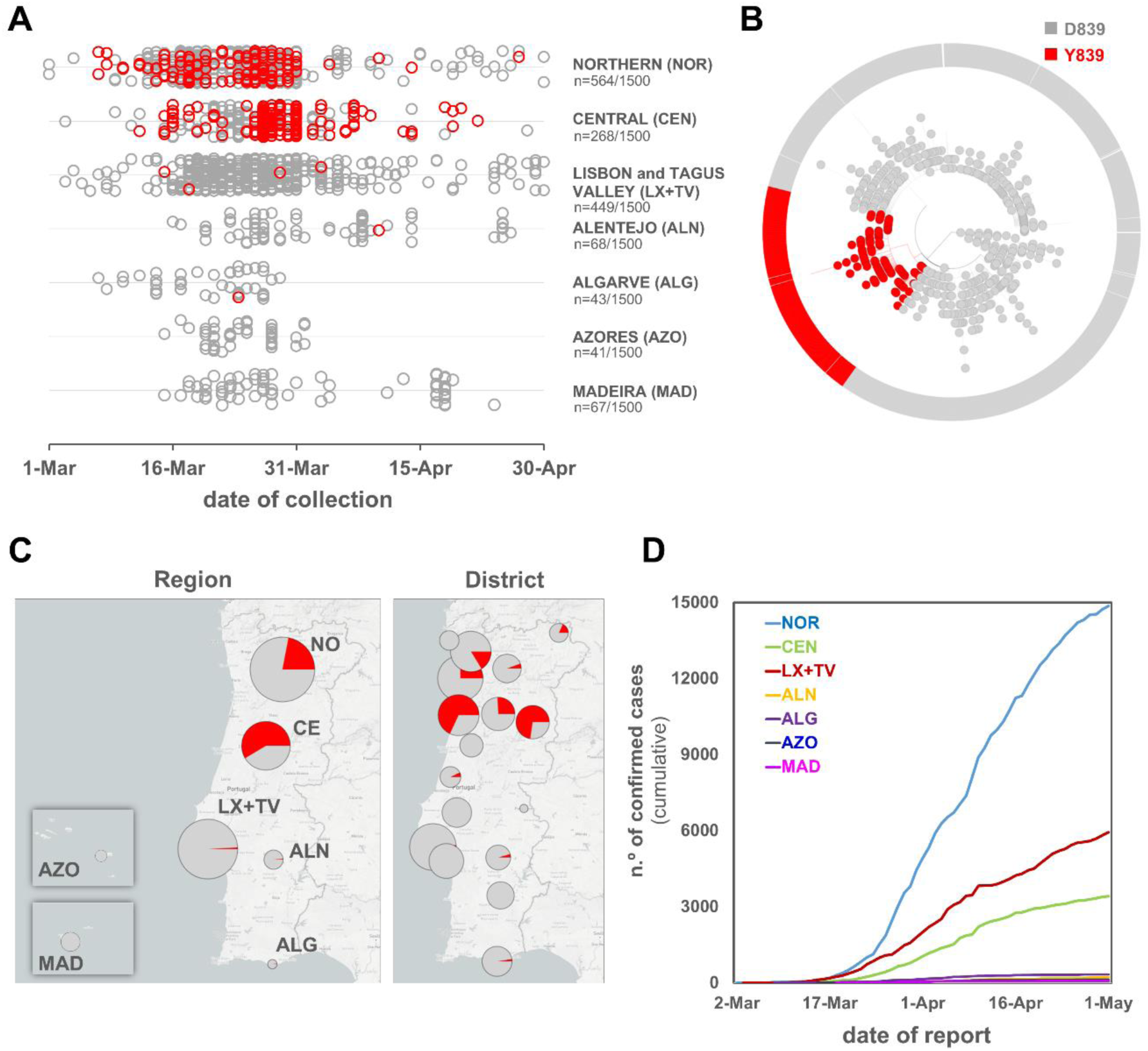
Landscape of the geotemporal spread of SARS-CoV-2 Spike Y839 variant in Portugal by region, as of April 30^th^’ 2020. **A** Distribution of the analysed genome sequences (n=1500) by date of sample collection and Health Administration region, highlighting COVID-19 cases caused by the Spike Y839 variant (red dots). **B**. Radial maximum likelihood phylogenetic tree showing the high proportion of genomes with the Spike D839Y mutation detected in Portugal [about 20% of all sequences collected until the end of March (255/1275) or the end of April (287/1500)]. This dataset covers 15.5% (1275/8251) and 6.0% (1500/24987) of all confirmed cases detected until the end of March and April, respectively. The phylogeny and geotemporal distribution can be visualized interactively at https://microreact.org/proiect/nDGsJKFv7gQTi1q8CQwwKR/0489f840 (geographic resolution by Region) and https://microreact.org/proiect/2kh3TRVYB9gWGRpNSJWDW5/b6c659e0 (geographic resolution by District) using Microreact (https://microreact.org/). **C**. Distribution of the Spike Y839 variant by Health Administration region, highlighting its high relative frequency in the Northern and Central regions of Portugal, where this variant represented 22% and 59% of the sampled genomes until the end of April 2020, respectively. The size of the pie charts is proportional to the number of sequenced genomes. **D**. Cumulative total number of COVID-19 confirmed cases by Health Administration region, showing the Northern region as the “epicenter” of the epidemic during the two first months (source: General Directorate of Health (DGS), https://covid19.min-saude.pt/relatorio-de-situacao/).

**Figure 3.**
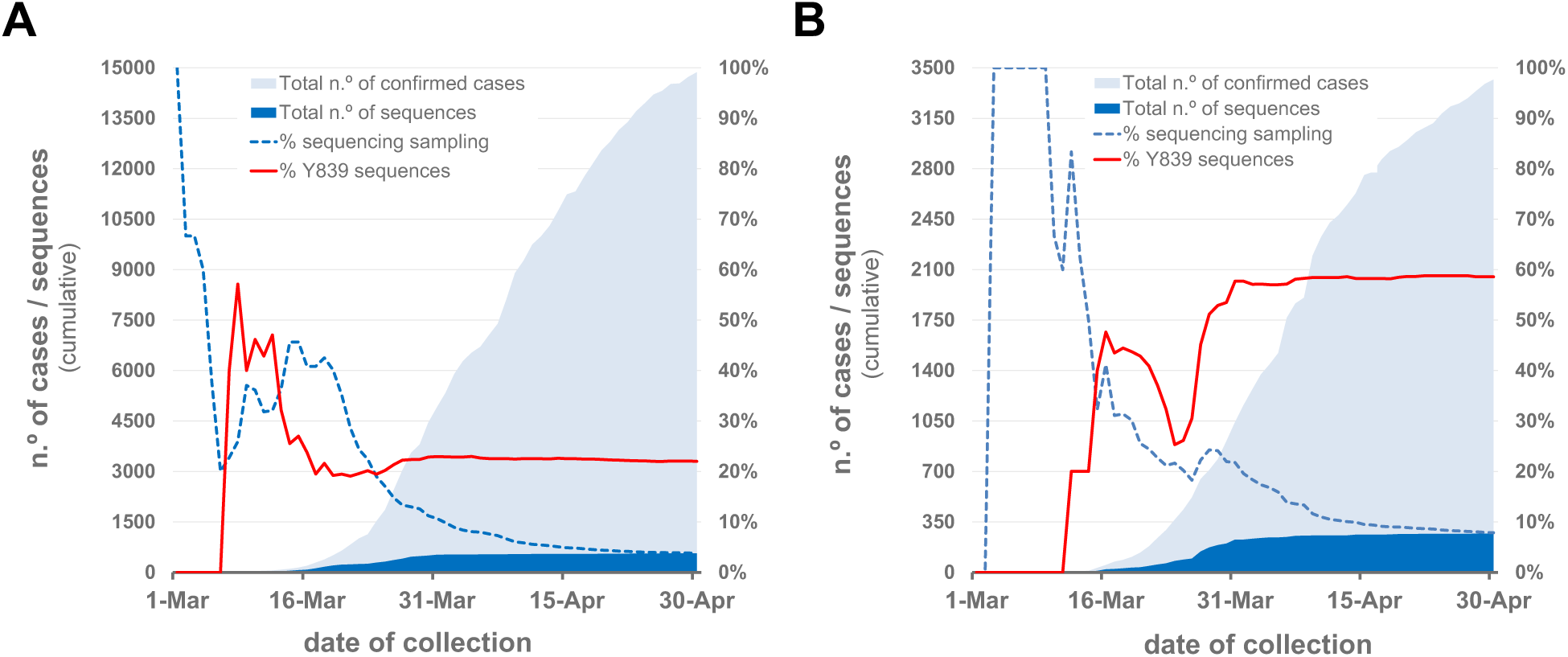
Overview of the SARS-CoV-2 genome sequencing sampling and cumulative relative frequency of the circulating Spike Y839 variant, as of April 30^th^, 2020 (n=1500), in the Northern (A) and Central (B) regions of Portugal. Area plots (left y-axis) reflect the cumulative total number of COVID-19 confirmed cases (light blue) and SARS-CoV-2 genome sequences (dark blue) detected/generated in each Health Administration region. Lines (right y-axis) display the cumulative percentage of COVID-19 confirmed cases with SARS-CoV-2 genome data, i.e., sequencing sampling (blue dash line) and the cumulative proportion of the Spike Y839 variant sequences (red line) detected in those regions during the same period.

**Figure 4.**
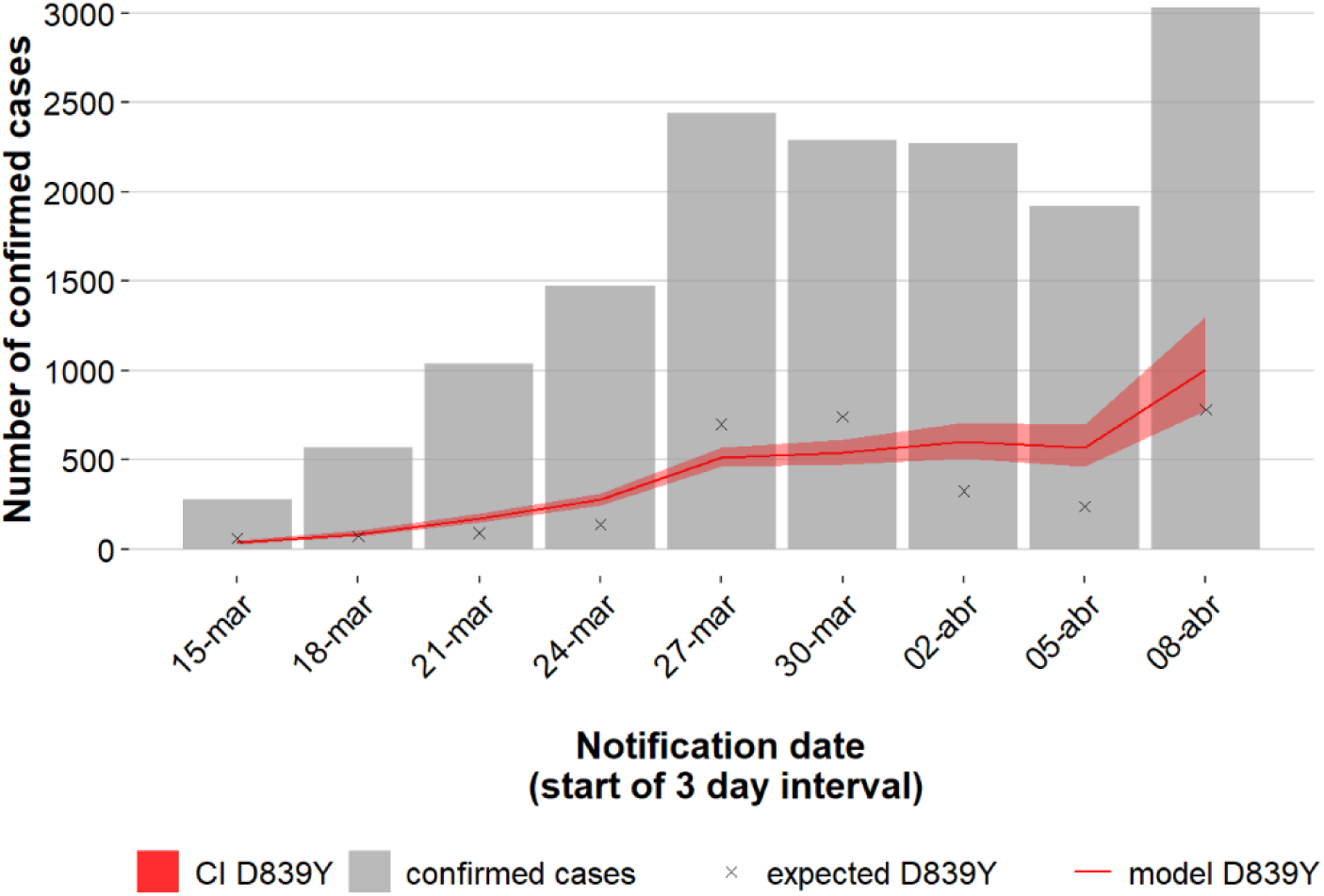
Increasing trajectory of the Spike Y839 variant and estimated weight of this variant in the total number of COVID-19 confirmed cases in early epidemic. Bar plot presents the total number of confirmed cases at each 3-day interval. A binomial regression model with logarithmic link function was applied to assess the temporal variation in the proportion of the Y839 variant among sequenced samples. This model was then applied to extrapolate the evolution of Y839 cases (red line) in the total case population (gray bars). Crosses represent the estimated Y839 cases and the shaded region shows the 95% confidence interval. 1 -day was assumed as the timeframe delay between sample collection and case notification.

### Detection and circulation of SARS-CoV-2 Spike 839 amino acid variants worldwide

To explore the frequency of SARS-CoV-2 D839Y mutation (and other mutations in 839 protein position) at worldwide level, we downloaded 66548 amino acid sequences (and associated metadata) of SARS-CoV-2 Spike protein available at GISAID (as of 23 July 2020). From the 65367 sequences collected outside Portugal having data for the 839 position, 97 were found to display mutations in this amino acid of interest (Table 1, Table S2). Of those, 92 genomes revealed the same genetic signature to that found in the 290 genomes from Portugal, i.e., a G-to-T nucleotide substitution in 24077 genome position (leading to the D839Y amino acid replacement) in a SARS-CoV-2 with Spike G614 background. As previously noticed (Korber et al, 2020), this data sustains that the global frequency of amino acid variants in this 839 site remains around 0.5% (similar estimates can be found in https://cov.lanl.gov/content/index and https://bigd.big.ac.cn/ncov/). After the first Spike Y839 variant was detected in Italy (Lombardy) on February 21^st^, the mutation D839Y has been reported in 12 other countries from four continents (Europe, Oceania, Asia and America) (Figure 5). Despite the highly unequal sequencing "sampling” (i.e., proportion of confirmed cases with SARS-CoV-2 genome data) between the different countries, it is noteworthy that, apart from Portugal, D839Y genomes represent >5% of all sequences made available at GISAID (as of July 23^rd^, 2020) by three countries (Estonia, Georgia and New Zealand) (Figure 5). Of note, the four genomes detected in Iceland on early March are also associated with travel history to Italy (Gudbiartsson et al, 2020; Nextstrain) (Table S2), consolidating our hypothesis about Spike Y839 variant “origins” in Italy.

**Table 1.**
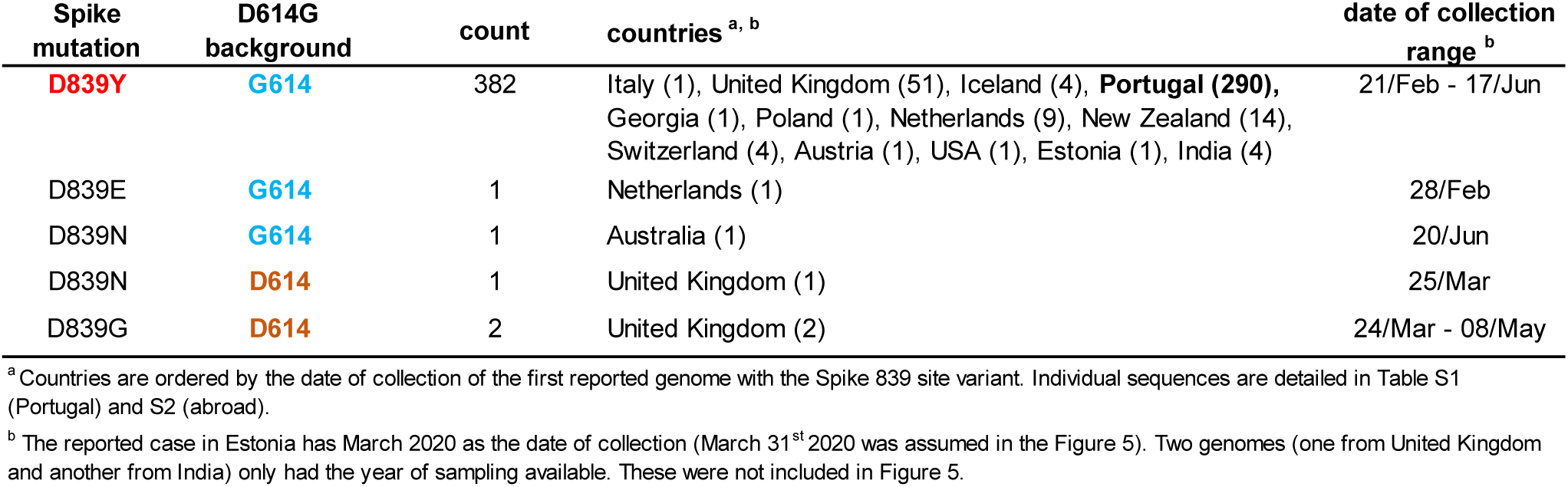
Overview of the SARS-CoV-2 Spike amino acid sequences with mutations in the 839 site available at GISAID, as of July 23^rd^, 2020.

**Figure 5.**
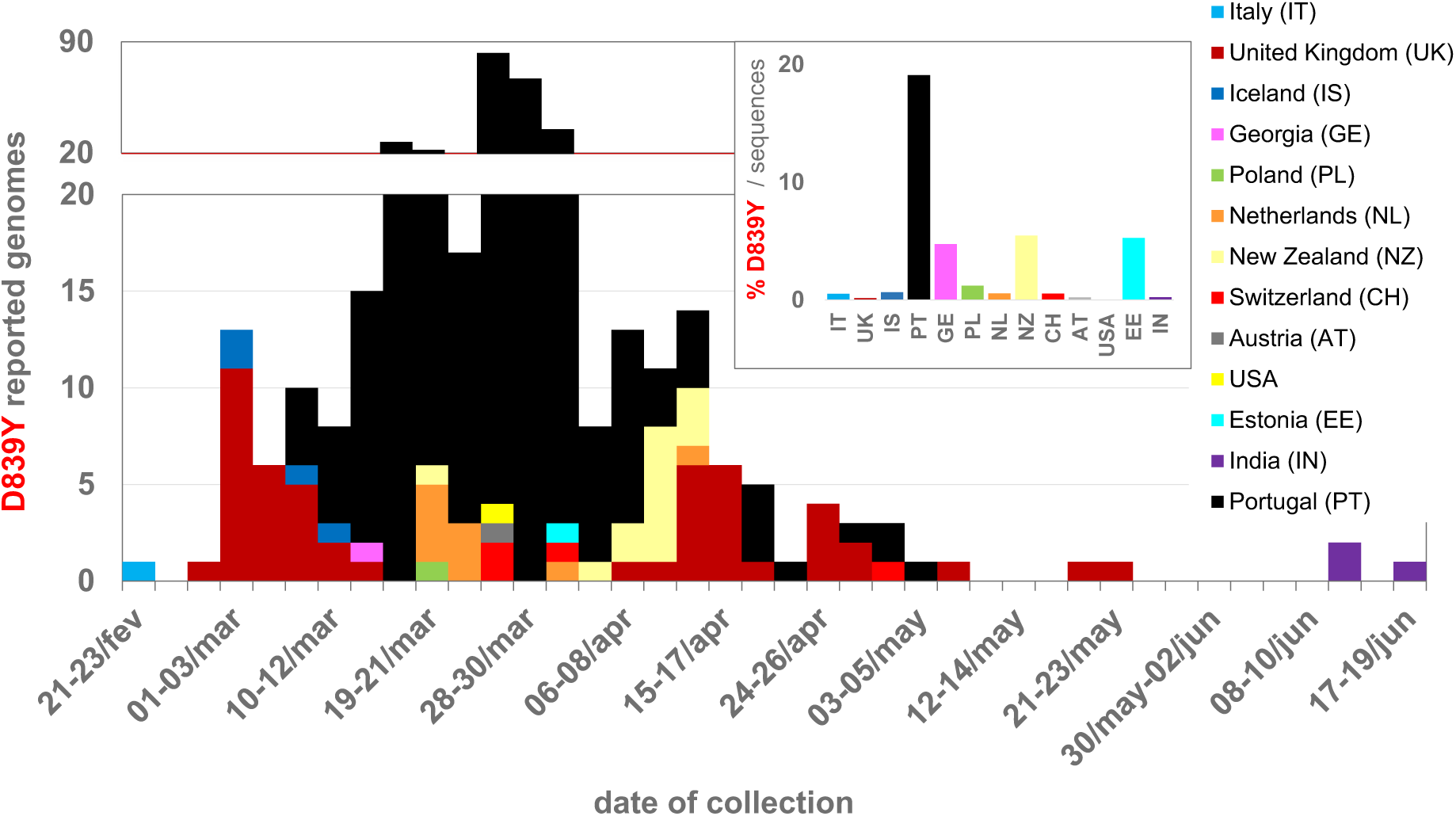
Detection and circulation of the SARS-CoV-2 Spike D839Y mutation worldwide. SARS-CoV-2 Spike amino acid sequences available at GISAID (https://www.gisaid.org/), as of July 23^rd^, 2020, were download, aligned and screened for the presence of mutations in Spike 839 amino acid position. The main plot display the country and data of collection of 92 Spike sequences with the D839Y mutation (detailed in Table S2). The bar graph in the upper right corner displays the proportion of D839Y sequences in the total number of Spike amino acid sequences available per country. As detailed in Table S2, the D839Y sequence from Estonia indicates March 2020 as the date of collection (March 31^st^ 2020 was assumed in this plot). Two genomes (one from United Kingdom and another from India) only had the year of sampling available, thus they were not included in the graph.

Notwithstanding, fine-tune integration of the 92 Spike Y839 genomes detected abroad in the “global” phylogeny (using Nextstrain https://nextstrain.org/ncov and Nextclade https://clades.nextstrain.org/) pointed out that the D839Y amino acid change likely emerged independently in two other instances. This observation is supported by one genome sequence collected in the United Kingdom (Wales) on April 24^th^ (Wales/PHWC-35B01/2020; GISAID accession EPI_ISL_474528), which presents all clade-defining SNPs of Nextstrain Clade 20C plus four additional SNPs (including G24077T). In this particular case, we cannot exclude the hypothesis that the G24077T nucleotide change might have been introduced in a clade “20C” SARS-CoV-2 by recombination, as “20C” and “20A harboring Y839” viruses co-circulated in Wales during the collection period (13 out of the 51 Spike 839Y genomes from UK were collected in Wales between April 10^th^ and May 25^th^). The potential third independent emergence of a Spike Y839 variant is supported by the four genomes collected in India, as they cluster apart from other G614+Y839 genomes, forming a subbranch (supported by 6 SNPs, including G24077T) within a large cluster mostly enrolling genomes from India. Again, the very limited sequencing sampling in some countries hampers a more in-depth assessment of Y839 origins, spread and global prevalence.

### Impact of Spike Y839 variant on viral load

The Cycle threshold (Ct) obtained in diagnostic PCR is an indirect indicator for relative viral loads in vivo, with lower Ct values indicating higher viral loads. Hence, when a given variant raises in frequency in the population and/or is potentially linked to more severe disease, there is an expected interest in assessing whether such variant is associated with lower or higher Ct values than its ancestral, as a possible indicator that the emergent mutation might have mediated modified transmissibility, infectivity or disease severity. Still, for SARS-CoV-2, increasing reports have been linking the globally dispersed Spike G614 variant to lower Ct values (Korber et al, 2020; Lorenzo-Redondo et al, 2020; Wagner et al, 2020), suggestive of higher upper respiratory tract viral loads leading to increased G614 transmissibility, but not pathogenicity (Korber et al, 2020).

As the National Reference Laboratory, INSA could gather Ct values associated with 940 out of the 1516 genomes analyzed in present study. Despite the expected bias associated with sample collection (e.g., momentum), sampling (e.g., selection) and testing (e.g., extraction, PCR protocol and equipment) within datasets enrolling multiple laboratories, we sought to verify whether the same trend is observed for the D614/G614 comparison and whether the Spike Y839 variant (Spike background G614+Y839) may also be potentially linked with changes in Ct values (viral loads). As previously seen (Korber et al, 2020; Lorenzo-Redondo et al, 2020; Wagner et al, 2020), we observed that infected individuals with Spike G614 had Ct values (mean=22.2; n=844) lower than those infected with D614 viruses (mean=22.9; n=96), still without statistical significance by Wilcoxon Rank Sum Test (Figure S4). Considering the phylogeny of SARS-CoV-2 in Portugal, we then compared Ct values according to D614/G614 status, phylogenetic group within G614 (i.e., 20B or non-20B) and Spike D839/Y839 status (Figure S4; https://microreact.org/proiect/nDGsJKFv7gQTi1q8CQwwKR/f46f1fa4). Curiously, Nextstrain “20B” clade (marked by the GGA-to-AAC SNP triplet at genome position 28881-3) revealed the lowest Ct values (mean=21.7; n=413) among all groups, although no statistical significance was observed in pairwise comparisons. Regarding the D839/Y839 comparison, we observed that Y839 variant presented non-significant higher Ct values (mean=22.7; n=220) than the ancestral D839 (mean=22.1; n=720). However, considering that D839 includes both 20B and non-20B samples, contrarily to Y839, which is a sub-group within 20A (Figure S4), we repeated the analysis by excluding 20B samples from the comparison and observed similar average Ct values in D839 (mean = 22.6; n=624) and Y839 (mean = 22.7; n=220) groups. When applying the same rationale (i.e., excluding 20B) to the D614/G614 comparison, the 0.7 difference observed in the mean Ct values using whole dataset decreased to less than 0.3.

## Discussion

One of the main objectives of conducting genome-based surveillance of circulating pathogens is to identify mutations potentially leading to fitness advantages and/or immunological / drug resistance. For SARS-CoV-2, special focus has been given to identify and monitor mutations affecting antigenic and other host-interacting regions, as well as mutations potentially associated with frequency sweeps at local, regional or global levels (Nextstrain; GISAID; Korber et al, 2020). In this perspective, mutations in the SARS-CoV-2 Spike D839 site, which is predicted to fall within the Spike fusion peptide (Tang et al, 2020; Korber et al, 2020), are of particular interest given both its non-negligible relative frequency worldwide and the pivotal role of this protein motif in inserting SARS-CoV-2 into human cell membranes Tang et al, 2020; Korber et al, 2020). In the present study, by tracking the temporal and geographical spread of a SARS-CoV-2 variant with a Spike D839Y amino acid change in Portugal, we show that this variant, which was likely originated from a single introduction from Italy in mid-late February 2020, was responsible for the largest SARS-CoV-2 transmission chain during the early epidemics, likely causing 24.8% (20.8-29.7%, CI 95%) COVID-19 cases in Portugal during the same period. Our data shows that it was circulating in Portugal before the first COVID-19 confirmed case was detected on March 2^nd^, becoming notably prevalent in the Northern and Central regions of the country, representing 22% and 59% of the sampled genomes until the end of April 2020, respectively (Figures 2 and 3). The Spike Y839 variant was introduced in the Northern region (the epicenter of the COVID-19 epidemic during the first two months), it was strongly linked to a massive cluster of cases in a coastal municipality (Ovar) from the Central region (the only municipality subjected to strict local quarantine) and progressively disseminated to inland locations of the Central region (Figure 2, S1 and S3).

Two major explanations can be drawn to justify the high prevalence of the Spike Y839 variant in Portugal. On the one hand, its high prevalence might be due to a populational/epidemiological effect (founder effect) as this variant was likely one of the first SARS-CoV-2 lineages introduced in Portugal. In this hypothesis, the increasing frequency trajectory of Spike Y839 variant would not have been mitigated because this variant disseminated well before the first confirmed cases in Portugal, when contact tracing, broad testing, and strict lockdown measures were still not in place. For example, on March 9^th^, case definition for a COVID-19 suspected case did not include individuals with acute respiratory infection, unless they required hospitalization, reported travel history or any contact with suspected or confirmed cases in the 14 days before symptoms onset (https://www.dgs.pt/directrizes-da-dgs/orientacoes-e-circulares-informativas/orientacao-n-002a2020-de-25012020-atualizada-a-250220201.aspx). On the other hand, the high prevalence could have been driven by fitness increase mediated by D839Y mutation. In this hypothesis, this mutation would have posed “advantageous” structural changes in the Spike protein with potential impact on SARS-CoV-2 infectivity and consequently, on its transmissibility, as suggested for D614G (Korber et al, 2020). While D614G was hypothesized to increase SARS-CoV-2 infectivity by influencing the dynamics of the spatially proximal fusion peptide (Korber et al, 2020), D839Y, which itself directly changes the fusion peptide (Tang et al, 2020; Korber et al, 2020), could also have shaped this motif towards a better fitted fusion of SARS-CoV-2 with human cells. Also, a recent preprint report (not peer reviewed) using computational modelling suggested that mutations in Spike D839 [in particular, to an aromatic tyrosine (Y)] may strengthen the interaction between the virus and human T cells, potentially influencing host inflammatory responses (Cheng et al, 2020). Although these clues about the potential functional effects of D839Y require more evidence through experimental validation, they clearly point out that a more in-depth study of the potential impact of this mutation of interest on SARS-CoV-2 transmissibility/pathogenicity should be conducted. In the same perspective, although we did not observe any association between Y839 and Ct values (viral load) in our dataset, this assessment needs to be revisited as more data is acquired. Notwithstanding, we indirectly observed that samples from the 20B clade (marked by the GGA-to-AAC SNP triplet at genome positions 28881-3) had a lower Ct than non-20B groups in our dataset, supporting that it is worth performing this comparison with additional datasets. Our observations sustain that it is challenging to discern the significance of particular mutations alone from population genetics, and that the evaluation of (sub)clade effects should not be neglected in this kind of screenings.

It is plausible that both hypotheses raised above (founder effect *versus* selective advantage) might have concomitantly contributed to the high frequency of the Spike D839Y mutation in Portugal. Its detection in 13 countries from four continents, its probable independent emergence in distinct times and genetic clades (20A and 20C) in some of these countries and its considerable relative weight (~5%) in the sampled genomes of three countries (besides Portugal) seem to favor the selective advantage hypothesis. Contrarily to Spike G614 variant, which emerged way before the general quarantine in Europe, the Spike Y839 variant likely emerged on mid-late February in Italy, when rigid lockdown measures started being implemented everywhere in Europe. This posed strong bottlenecks on SARS-CoV-2 population, giving Y839 variant less opportunity to expand, even if it was selectively advantageous over other circulating variants. Still, the founder effect and timeline of the epidemics also certainly favored its remarkable increase in frequency in Portugal, contrarily to what might have happened in other countries. For instance, first lockdowns in Lombardy, Italy, where Spike Y839 variant was firstly detected on February 21^st^, coincidently began on this date (https://en.wikipedia.org/wiki/COVID-19 pandemic lockdown in Italy). In Portugal, national quarantine was implemented on March 18^th^, when Spike Y839 variant had already been circulating in the community for at least three weeks. Nonetheless, it is worth highlighting that the huge discrepancies in sequencing sampling between countries completely hampers a real knowledge of D839Y frequency regionally and globally. Our sequencing sampling after April 30^th^ does not allow us to infer the current relative frequency of the Spike Y839 variant in Portugal. One can speculate that its circulation was highly contained considering that, after this period, the epidemic evolved favorably in Northern and Central regions (where Y839 had mostly circulated), contrarily to the Lisbon and Tagus Valley region (where Y839was, at that time, rarely seen due to the confinement measures in the North and Central regions) (INSA, COVID-19 epidemic evolution, August 7^th^, 2020; http://www.insa.min-saude.pt/wp-content/uploads/2020/08/Report_covid19_07_08_2020.pdf).

In summary, we describe the emergence and increase in frequency of a Spike Y839 variant that reached a tremendous impact on COVID-19 epidemic in Portugal, as estimated by its high relative weight of one in each four cases during the exponential phase of the epidemic. Our study reinforces the need for continuous and close surveillance of SARS-CoV-2 genetic diversity, with emphasis on detecting and monitoring variants with potential impact at biological and/or epidemiological levels, as a result, for instance, of increased fitness or immunological/drug resistance.

## Material and Methods

### Sample collection

Samples used in this study were collected as part of the ongoing national SARS-CoV-2 laboratory surveillance conducted by the National Institute of Health (INSA) Doutor Ricardo Jorge, Portugal. INSA receives both clinical specimens for SARS-CoV-2 molecular testing and confirmation by the National Influenza Reference Laboratory and other Respiratory Viruses (designated as the National Reference Laboratory providing confirmatory testing for SARS-CoV-2) and SARS-CoV-2 positive samples (either clinical specimens or RNA) from a nationwide network with >50 laboratories that was rapidly established during the start of COVID-19 pandemics (here called “Portuguese network for SARS-CoV-2 genomics”). Demographic information, date of sample collection, date of illness onset and travel history were obtained when possible. Geographical data presented in this study refers to the Region (“Health Administration region”), District or Municipality of the patients’ residence or, when no information is available (for a small proportion of cases), to the location of exposure or of the hospital/laboratory that collected/sent the sample.

### SARS-CoV-2 genome sequencing

SARS-CoV-2 positive RNA samples were subjected to genome sequencing using a whole-genome amplification strategy with tiled, multiplexed primers (Quick et al, 2017) and the Artic Consortium protocol (https://artic.network/ncov-2019; https://www.protocols.io/view/ncov-2019-sequencing-protocol-bbmuik6w), with slight modifications. In brief, after cDNA synthesis using SuperScript™ IV First-Strand Synthesis System kit (Invitrogen™, catalog: 18091050) with random hexamers from 11 μL of RNA (exactly as described in Artic Network protocol), targeted amplification was performed with 2.5 μL of cDNA using NEBNExt® Q5® HotStart HiFi Master Mix (12.5 μL per reaction) (New England BioLabs, catalog: M0544S) with two pools of tiling primers (A and B) separately. Primers versions V1 and V2 (aliquots kindly provided by Artic Network team) were used for the first 243 samples of this study, while the V3 primers (with a total of 218 primers) were applied to all samples afterwards (all versions available here: (https://github.com/artic-network/artic-ncov2019/tree/master/primer_schemes/nCoV-2019). The final concentration per primer (V3 version) was ~0.013μM (1.4 μM per pool) in a 25 μL total reaction volume. PCR amplification parameters were: 30s at 98°C, 35 cycles of 15s at 98°C and 5 min at 65°C (for the first 762 samples) or 63°C (afterwards), and final extension for 5 min at 65/63°C. Amplicons were visualized on a 1% agarose gel, tubes A and B were pooled per sample, and subjected to clean up with Agencourt AMPure XP (Beckman Coulter, catalog: A63880) using a 1:1 volume ratio. Dual-indexed sequencing libraries were prepared using Illumina Nextera XT DNA Library Prep Kit (Illumina). Pooling, denaturation and dilution of bead-based normalized libraries was performed according to the manufacturer’s instructions for the MiSeq or NextSeq 550 systems (Illumina). A 1% spike-in PhiX genome library (Illumina) was used as internal quality control. Libraries were sequenced using 250bp (MiSeq) or 150bp (NextSeq 550) paired-end reads targeting ~1M reads per sample.

### Genome assembly and sequence curation

Analysis of sequence read data was conducted using the bioinformatics pipeline implemented in INSaFLU (https://insaflu.insa.pt/; https://github.com/INSaFLU), which is a web-based (and also locally installable) platform for amplicon-based next-generation sequencing data analysis (Borges et al, 2018). Briefly, the core bioinformatics steps (documented in Borges et al, 2018 and https://insaflu.readthedocs.io/) involved: i) raw NGS reads quality analysis and improvement using FastQC; (https://www.bioinformatics.babraham.ac.uk/projects/fastqc) and Trimmomatic (http://www.usadellab.org/cms/index.php?page=trimmomatic), respectively (read’s ends were cropped 30bp for primer clipping); ii) draft *de novo* assembly using SPAdes (http://cab.spbu.ru/software/spades/) followed by classification and contigs assignment of Human Betacoronavirus; and, iii) reference-based mapping, consensus generation and variant detection using the multisoftware tool Snippy (https://github.com/tseemann/snippy), using the Wuhan-Hu-1/2019 genome sequence (https://www.ncbi.nlm.nih.gov/nuccore/MN908947) as reference. The first 40bp and end 100bp were discarded and consensus sequences were exclusively included in the study when >70% of genome was covered by at least 10-fold. For samples with coverage drop below 10fold, fine-tuned consensus sequence curation was performed as follows: i) undefined bases (“N”) were placed in genome regions with depth of coverage below 10 using a python script (https://github.com/rfm-targa/BioinfUtils/blob/master/msa_masker.py), and “N” regions at the sequence ends were trimmed to avoid releasing sequences starting or ending with “N”; ii) all regions with depth coverage below 10 were visually inspected in the Integrative Genomics Viewer (http://www.broadinstitute.org/igv) available *in situ* at INSaFLU, and when mutations were detected and validated within these low coverage regions, they were inserted in the consensus sequence to avoid disrupting/biasing the phylogenetic signal; iii) when a mutation was validated within a “N” region with <=100bp, the whole region was inserted (as long as the region was covered by at least one read); when a mutation was validated within a “N” region with >100bp, the mutation was inserted together with additional 20 bp (10bp from each flanking side) to improve the downstream sequence alignment.

### Phylogenetic analysis and real-time data sharing on SARS-CoV-2 genetic diversity and geotemporal spread in Portugal

A total of 1516 SARS-CoV-2 genome sequences were analyzed in this study (corresponding to INSA’s collection as of July 23^rd^, 2020; Table S1) using the SARS-CoV-2 Nextstrain pipeline (Hadfield et al, 2018) version from March 23, 2020 (https://github.com/nextstrain/ncov), with slight modifications. In brief, sequences were aligned against the reference Wuhan-Hu-1/2019 genome of SARS-CoV-2 (GenBank accession MN908947) using MAFFT (Kaoth et al, 2002), the alignment was visually inspected, manually curated and further used to build a maximum likelihood phylogenetic tree based on the GTR model using IQ-TREE (Nguyen et al, 2015) following the Nextstrain implementation (the first 130bp and last 50 bp, as well as a few bases within the alignment, were masked as likely sequencing artifacts). Within the Nextstrain pipeline, Treetime (Sagulenko et al, 2020) is applied to infer a time-resolved phylogeny. The phylogeny is rooted relative to early samples from Wuhan, China (Wuhan-Hu-1/2019, GenBank accession MN908947; Wuhan/WH01/2019, GenBank accession LR757998) and temporal resolution assumes a nucleotide substitution rate of 0.0008±0.0004 substitutions *per* site *per* year.

A website (https://insaflu.insa.pt/covid19) was launched on March 28, 2020 for real-time data sharing on SARS-CoV-2 genetic diversity and geotemporal spread in Portugal. This site gives access to “situation” reports of the study and provides interactive data navigation using both Nextstrain (https://nextstrain.org/) (Hadfield et al, 2018) and Microreact (https://microreact.org/) (Argimón et al, 2016) tools. As of July 23, 2020, “clade" assignment reflects the Nextstrain classification (https://github.com/nextstrain/ncov; version June 3, 2020), while "Lineage" refers to the classification based on Phylogenetic Assignment of Named Global Outbreak Lineages (Pangolin) (https://github.com/hCoV-2019/pangolin - lineage_version: 2020-05-07) (Rambaut et al, 2020).

In this study, both Nextstrain (https://nextstrain.org/) (Hadfield et al, 2018) and Microreact (https://microreact.org/) (Argimón et al, 2016) visualization tools were used to deeply explore the genetic diversity and geotemporal spread dynamics of SARS-CoV-2 Spike D839Y variant in Portugal. The phylogeny and associated metadata used in this study can be visualized interactively at https://microreact.orq/proiect/nDGsJKFv7qQTi1q8CQwwKR/18a0a470 (global dataset, as of July 23^rd^, 2020), https://microreact.org/proiect/nDGsJKFv7gQTi1q8CQwwKR/0489f840 (geographic resolution by Region, genomes collected until April 30^th^ highlighted) and https://microreact.org/proiect/2kh3TRVYB9gWGRpNSJWDW5/b6c659e0 (geographic resolution by District, genomes collected until April 30^th^ highlighted)) using Microreact (https://microreact.org/) (Argimón et al, 2016). To explore the frequency of SARS-CoV-2 Spike D839Y variant at worldwide level, we downloaded 66548 amino acid sequences (and associated metadata) of SARS-CoV-2 spike protein available at GISAID (as of 23 July 2020) and aligned them using MAFFT (Kaoth et al, 2002). A total of 65367 sequences collected outside Portugal had sequence data for amino acid position 839, giving a total of 66883 (65367 plus 1516) sequences screened in this study for the presence of mutations in this amino acid of interest. The 92 genomes sequences of the SARS-CoV-2 Spike D839Y variant detected abroad were downloaded from GISAD (GISAID acknowledgments are in Table S2). Their clade classification and integration into "global” and Portugal phylogeny was performed using Nextstrain (https://nextstrain.org/ncov) and Nextclade (https://clades.nextstrain.org/).

### Statistical analysis

In order to assess the temporal variation in the proportion of D839Y mutation among sequenced samples, a binomial regression model with logarithmic link function was applied. The model was then applied to extrapolate the evolution of Y839 cases in the total case population (data presented in Figure 4). To increase the robustness of the analysis, the studied timeframe (from March 14^th^ to April 9^th^ was adjusted to ensure 3-days bins with at least 25 genome sequences. This period overlaps the exponential phase of the epidemic in Portugal. We assumed one day as the timeframe delay between sample collection and case notification. The Kruskal-Walls non-parametric test was used to assess the existence of statistically significant differences in Ct values between groups (Figure S4). Differences in Ct values for each pair of groups were assessed using the Wilcoxon test adjusted for multiple comparison tests.

### Data availability

SARS-CoV-2 genome sequences generated in this study were uploaded to GISAID database (https://www.gisaid.org/). Accession numbers can be found in Supplementary material (Table S1).

### Ethical Approval

This study was approved by the Ethical Committee (“Comissao de Etica para a Saúde”) of the Portuguese National Institute of Health.

## Acknowledgments

We also gratefully acknowledge to Sara Hill and Nuno Faria (University of Oxford) and Joshua Quick and Nick Loman (University of Birmingham) for kindly providing us with some sets of Artic Network primers for NGS; Rafael Mamede (MRamirez team, IMM, Lisbon) for developing and sharing a bioinformatics script for sequence curation (https://github.com/rfm-targa/BioinfUtils); all authors, originating and submitting laboratories who have contributed genome data on GISAID (https://www.gisaid.org/) on which part of this research is based. This study is co-funded by Fundação para a Ciência e Tecnologia and Agência de Investigação Clínica e Inovação Biomédica (234_596874175) on behalf of the Research 4 COVID-19 call. This work is also a result of the GenomePT project (POCI-01-0145-FEDER-022184), supported by COMPETE 2020 - Operational Programme for Competitiveness and Internationalisation (POCI), Lisboa Portugal Regional Operational Programme (Lisboa2020), Algarve Portugal Regional Operational Programme (CRESC Algarve2020), under the PORTUGAL 2020 Partnership Agreement, through the European Regional Development Fund (ERDF), and by Fundação para a Ciência e a Tecnologia (FCT).

## Supplementary Figures

**Figure S1.**
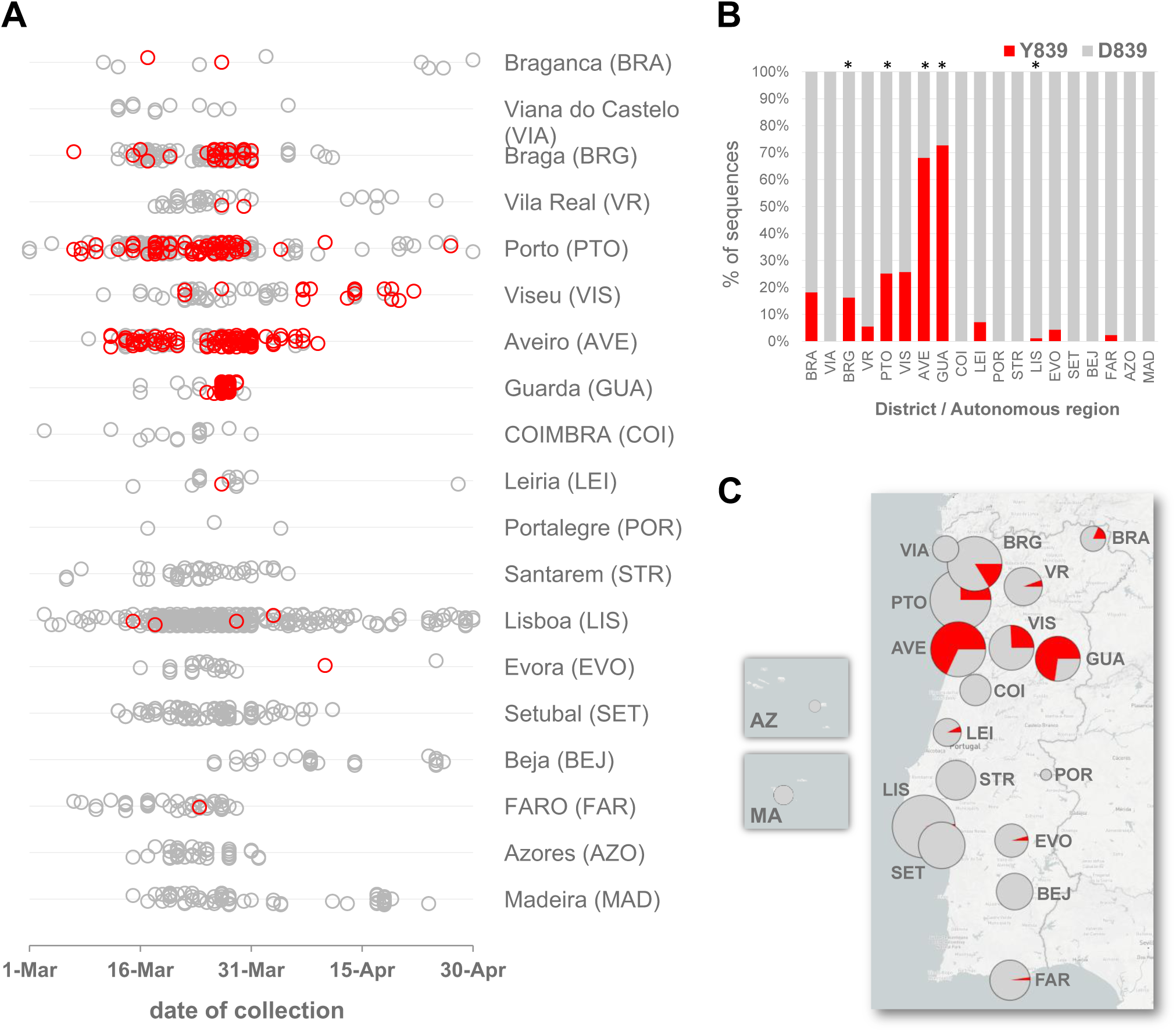
Landscape of the geotemporal spread of SARS-CoV-2 Spike Y839 variant in Portugal by District, as of April 30^th^’ 2020. **A** Distribution of the analysed genome sequences (n=1500) by date of sample collection and District, highligthing COVID-19 cases caused by the Spike Y839 variant (red dots). **B**. Relative frequency of the Spike Y839 variant across the 11 Districts where the variant was detected until April 30^th^. Asterisks above the graph denote Districts where more than 50 genomes were sampled. **C**. Relative frequency of Spike Y839 variant by District until the end of April 2020. The phylogeny and geotemporal distribution can be visualized interactively at https://microreact.org/project/nDGsJKFv7gQTj1q8CQwwKR/0489f840 (geographic resolution by Region) and https://microreact.org/project/2kh3TRVYB9gWGRpNSJWDW5/b6c659e0 (geographic resolution by District) using Microreact (https://microreact.org/).

**Figure S2.**
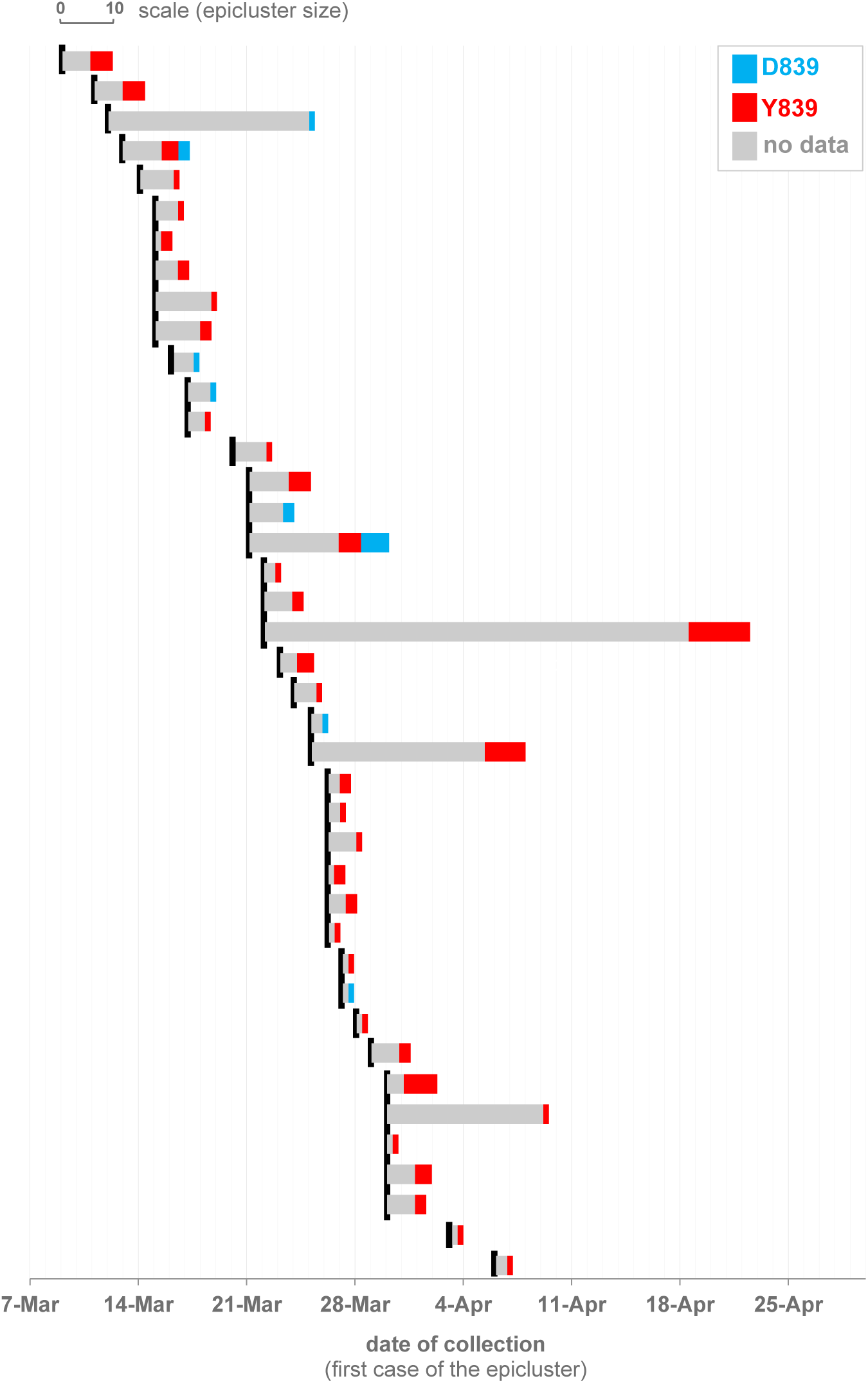
SARS-CoV-2 D839/Y839 status of epiclusters of potential epidemiologically linked confirmed cases (“epiclusters”) monitored by the Public Health Unit of Primary Care Cluster of Baixo Vouga, as of April 30^th^, 2020. The graph shows the temporal distribution (by date of sample collection of the first case of the epicluster; black lines) of 41 potential “epiclusters” (covering a total of 420 confirmed cases) for which SARS-CoV-2 genome data is already available. 33 epiclusters (323 confirmed cases) (77%) are exclusively associated with the Spike Y839 variant. The two potential epiclusters where both D839 and Y839 variants were ambiguously detected are under close contact tracing investigation to disclose this incongruence.

**Figure S3.**
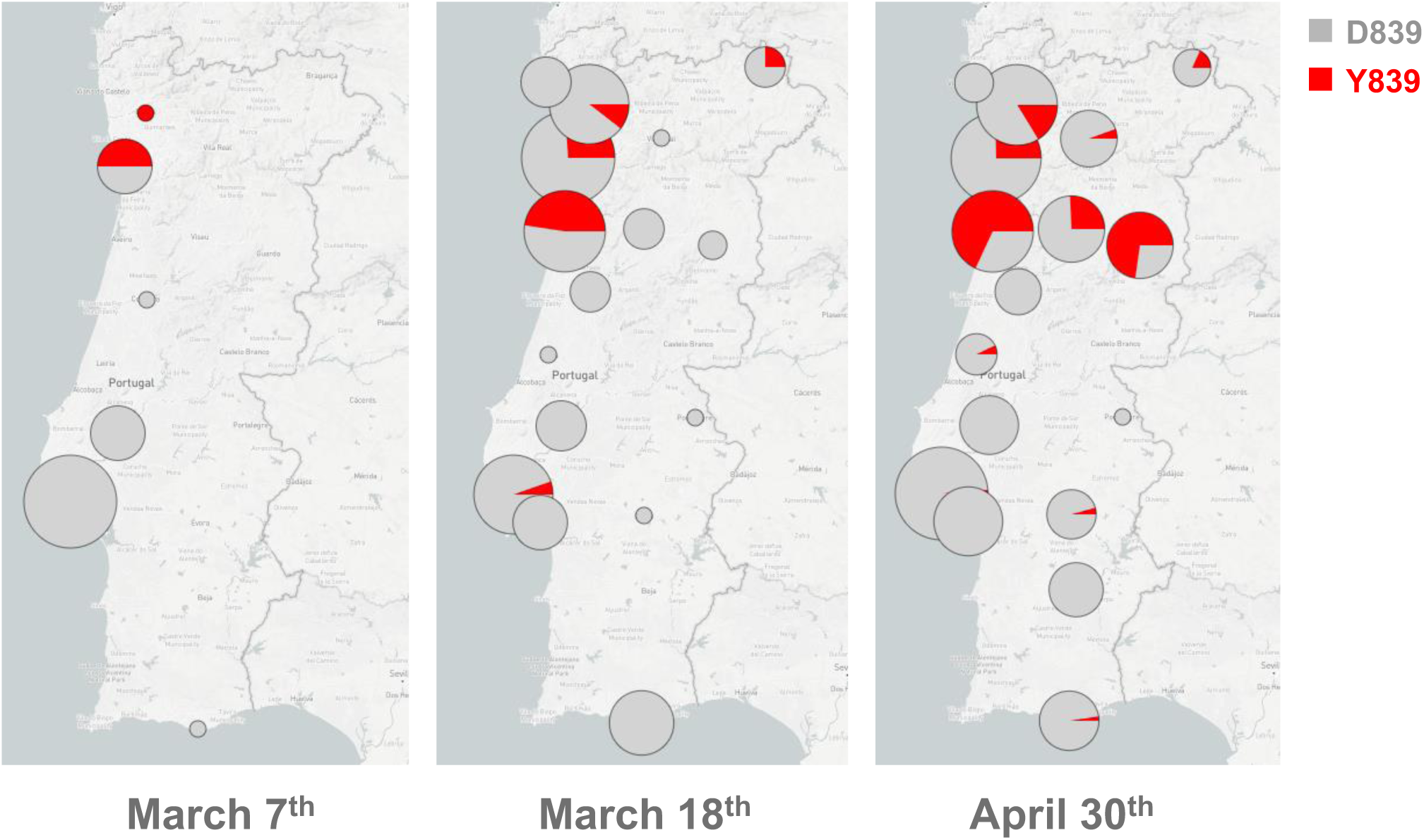
Geographical distribution of the relative frequencies of the D839 (gray) and Y839 (red) variants by District in three timeframes: March 7^th^ (when the first Y839 genomes were detected), March 18^th^ (when the emergency state was declared in Portugal and national lockdown was implemented) and April 30^th^’ 2020. Autonomous regions (Azores and Madeira) are not shown in these maps as no Y839 case where detected there (see Figure S1).The phylogeny and geotemporal distribution can be visualized interactively at https://microreact.org/project/nDGsJKFv7gQTj1q8CQwwKR/0489f840 (geographic resolution by Region) and https://microreact.org/project/2kh3TRVYB9gWGRpNSJWDW5/b6c659e0 (geographic resolution by District) using Microreact (https://microreact.org/).

**Figure S4.**
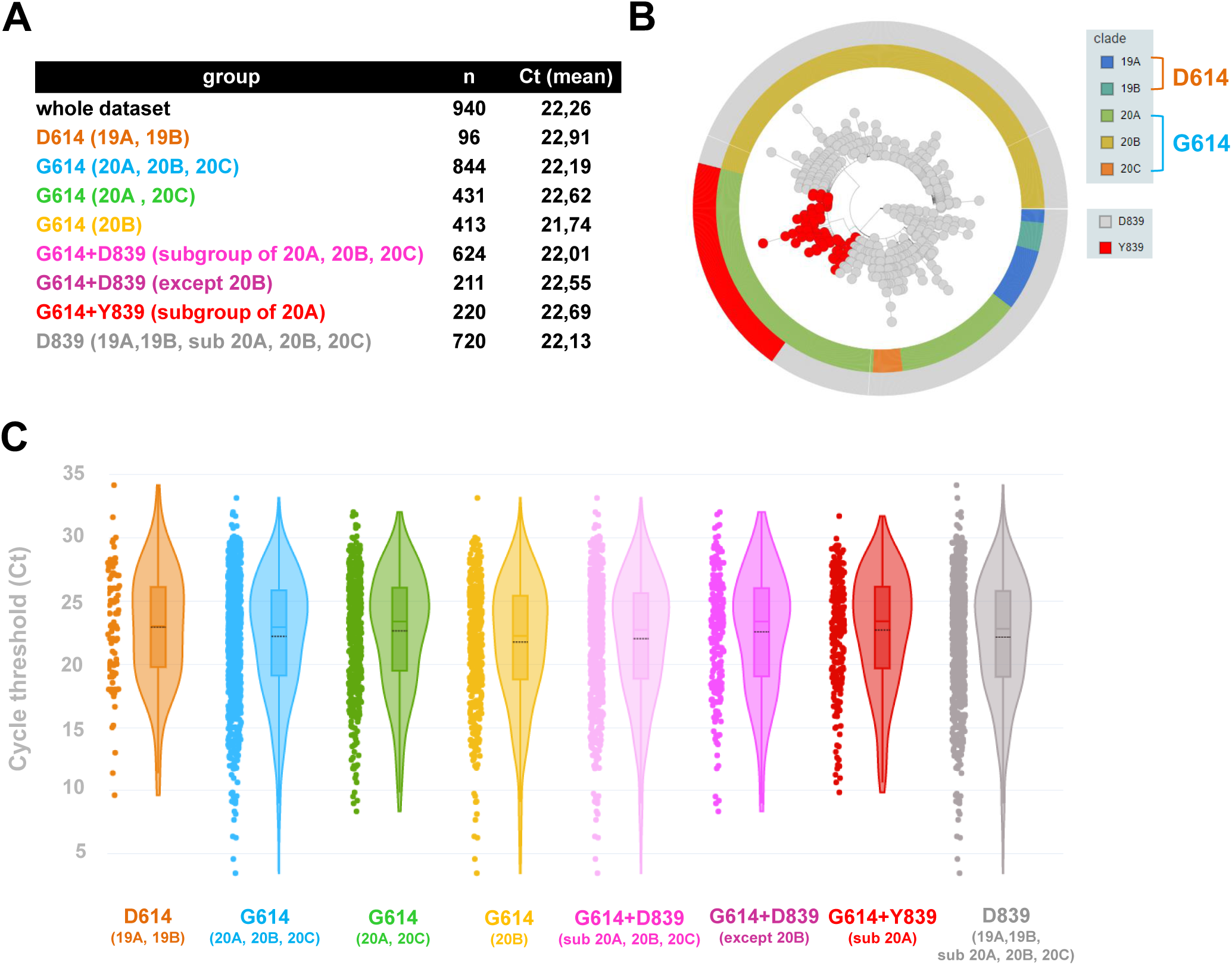
Comparison of Cycle threshold (Ct) values obtained in diagnostic PCR between different samples groups. Groups were established according to phylogenetic clustering and Nextstrain Clade classification of the genomes studied, D614/G614 status, phylogenetic group within G614 (i.e., 20B or non-20B) and Spike D839/Y839 status. **A**. Mean Ct values observed per group. **B**. Global phylogeny of the 1516 genomes studied (https://microreact.org/proiect/nDGsJKFv7gQTi1q8CQwwKR/f46f1fa4) highlighting the Nextstrain clades, D614/G614 and D839/Y839 status. **C**. Violin, scatter and box plots showing the dispersion of Ct values per group. Mean Ct values are indicated by black dash lines.

